# Development and validation of a generative AI-assisted medication-indication knowledge base

**DOI:** 10.64898/2026.01.06.26343341

**Authors:** Saniya Deshpande, Florence Ma, Alistair Marsland, Mattia Ficarelli, Linxuan Zhang, Alice Beattie, Edward Bonnett, Benjamin D Bray

## Abstract

**Background:** Existing information resources about medicines and their indications have limited usefulness for health data analytics. The emerging potential of large language models (LLMs) to generate clinically accurate responses presents a novel opportunity to develop a comprehensive knowledge base of medicines and their clinical indications.

**Method:** Unique medications from the English Prescribing Dataset (EPD) were extracted and included in a fine-tuned prompt pipeline using the GPT-4 and MedCAT LLMs. The resulting database underwent clinical validation by three clinicians to calculate the precision for a sample of the knowledge base. This was followed by external validation using the participant reported indications included in the National Health and Nutrition Examination Survey (NHANES) dataset, a large-scale population health survey in the United States (US).

**Results:** 1,540 unique medications from the EPD were used to generate 10,853 unique medication-indication pairs. Initial threshold-based investigations of accuracy across LLM generated confidence scores for each pair revealed that a threshold of 0.75 was an optimal trade-off between error rate and knowledge base size. Common types of error which had to be addressed in the pipeline included duplications, alternative spellings and medical synonyms. A random subset of 465 medication-indication pairs was selected for clinical validation, of which 418 were assessed to be correct (a precision of 89.9%). External validation using the NHANES overall agreement of 84% (210 out of 250). Of the remaining 40, only 2 were valid indications omitted in the knowledge base, whilst the rest were judged by clinical reviewers to be off-licence indications (n=7) or appeared to be incorrectly reported by survey participants (n=31).

**Conclusion:** The AI-assisted medication-indication knowledge base demonstrated high precision and external validity. The coverage of off-licence indications adds to the potential of this knowledge base in various biomedical applications such as real-world evidence research, drug discovery and adverse event detection.

## Background

Structured databases with information about medications and their clinical uses (“Indications”) have many applications in biomedical research. These include drug discovery and repurposing research, pharmacogenomic analysis, side effects identification and analysis and biomedical text mining^1^. Ideally, there would be one complete and actively maintained database which is kept up to date with new medicine launches and changes in clinical practice, is standardised to clinical terminologies and coding schemes (such as International Classification of Diseases, (ICD) and Systemized Nomenclature of Medicine(SNOMED)) and which would ensure consistency across studies and biomedical applications. However, such a dataset does not yet exist; many knowledge bases contain information extracted from drug labels and structured product leaflets (SPLs), so only focus on licensed indications (e.g., LabeledIn^2^., SIDER^3^, DICE^4^ and InContext^5^.), whilst others focus solely on off-licence indications^6^. Some are limited to only the most frequently prescribed medicines^2^ or are not available in a machine-readable format suitable for integrating into data analytics workflows (e.g., the British National Formulary, BNF^7^).

Creating and maintaining such a database requires expert clinical and pharmaceutical knowledge, making it very challenging to develop a database across tens of thousands of potential medicines-indications pairs. Recently several large language models have demonstrated clinical knowledge and reasoning abilities approaching and, in some studies, exceeding those of human physicians^8^. We therefore aimed to develop and validate a large language model (LLM) assisted medication-indication knowledge base, using the generative abilities of these models to overcome issues relating to scalability, whilst also compiling indication information from a much wider pool of data than drug labels or SPLs alone to improve coverage of the knowledge base.

## Methods

### Data sources

All medicines dispensed in the community in the United Kingdom were identified from the English Prescribing Dataset, which offers a comprehensive and unified source of information on prescriptions issued in England and dispensed across England, Wales, Scotland, Guernsey, Alderney, Jersey, and the Isle of Man^9^.

For external validation of the medication-indication database, the prescriptions drugs dataset from the National Health and Nutrition Examination Survey (NHANES)^10^ was used (2017-March 2020, P_RXQ_RX). NHANES is a programme of studies from the National Center for Health Statistics (NCHS), which is part of the Centers for Disease Control and Prevention (CDC). It uses interviews/questionnaires along with physical examinations and laboratory tests to assess the health and nutrition status of the US population. The prescriptions dataset is part of the questionnaire data of NHANES and includes patient-reported use of prescription medications along with the ICD-10-CM indication that most closely matches the reason for use.

### Prompt engineering and knowledge base creation

We employed OpenAI’s GPT-4 large language model^11^ via the OpenAI Application Programming Interface (API) to generate indications for each given medicine at the chemical substance level in our list. The model was accessed in April 2023, using the base GPT-4 model, with no plugins or fine-tuning. We iteratively developed the prompt instructions through a pilot testing phase. An initial prompt was drafted to instruct GPT-4 to produce a comprehensive list of all clinical indications for a given active ingredient (n = 1,540, at the chemical substance level) explicitly asking it include both licensed and off-licence uses. The prompt also asked the model to provide additional fields for each indication: the purpose of the medication (whether it is used for treatment or prevention), and a model-generated confidence score between 0 and 1. The confidence score was a self-reported measure output by GPT-4 reflecting its estimated confidence in the correctness of that medication-indication pair. We note that this confidence value is a heuristic provided by the model rather than a probabilistic guarantee – it offers a relative ranking of the model’s certainty for the indications but may not be perfectly calibrated. We used these confidence scores as a guide in subsequent filtering and validation steps (with the recognition of their limitations). This draft prompt was tested on a sample of common medications (e.g. atorvastatin, omeprazole, amlodipine; see Appendix for example) and refined in an iterative manner. We adjusted wording and format to ensure the model’s output was structured as a table and included all requested details without omissions. The finalised prompt (provided in the Appendix) instructed GPT-4 to return the medication-indication list in a JSON format for easy parsing. For each medication, we generated outputs from GPT-4 using two different randomisation settings to maximise recall of possible indications. Specifically, we invoked the model twice: once with a temperature setting of 0 (fully deterministic output) and once with a temperature of 0.2 (introducing slight variability). All other generation parameters were left at OpenAI’s default values (top_p = 1.0, frequency_penalty = 0, presence_penalty = 0). Using a temperature of 0 ensured that the most likely indications according to the model would consistently appear, while a temperature of 0.2 allowed the model to occasionally include fewer common indications. The outputs from these two runs were combined, and duplicate entries were removed, to form the initial version of the medication–indication knowledge base.

### Data cleaning and harmonisation

We implemented a post-processing pipeline to clean and standardise the GPT-4 outputs before validation. First, we removed any duplicate medication–indication entries following the merging of the two GPT-4 runs with different temperature settings. We then corrected obvious spelling mistakes and normalised terminology (for example, converting American spelling or terms to British English, such as “tumor” to “tumour” and “acetaminophen” to “paracetamol”). We also harmonised synonyms and variant phrasings for indications to ensure consistency. For instance, if the model output included both “atopic dermatitis” and “eczema” for a drug, these were recognised as the same indication; one term was chosen as the canonical representation to avoid duplicate entries.

### Data linkages

To enhance the interoperability of the knowledge base with other datasets and coding systems, we conducted additional linkages for each medication and indication. We performed approximate string matching to align each active ingredient to its corresponding standard code and description in the British National Formulary (BNF)^7^. We used the RecordLinkage package in R^12^ to compute Levenshtein string similarity between each GPT-4 medication name and the list of BNF medication names. A high similarity threshold was applied to identify the best match for each drug, and each proposed match was then manually verified to ensure the correct mapping. This step resolved minor naming differences (e.g. presence of salts, hyphens, or abbreviations) between the GPT-generated name and the official BNF name. After this harmonisation, each active ingredient in the knowledge base was linked to a BNF Chemical substance code, a standard identifier for medicines in UK prescribing data.

We then mapped each indication output by GPT-4 to clinical terminology codes. We applied MedCAT (Medical Concept Annotation Toolkit)^13^– a natural language model trained on hospital electronic medical records, specifically designed for natural language processing of clinical text. MedCAT was used to identify and map each indication to SNOMED CT concepts and, where possible, to ICD-10 diagnostic codes. This provided a consistent coding for the condition names, handling cases where different terms referred to the same underlying concept. The use of MedCAT helped to resolve ambiguous terms and ensured that indications were expressed in standard clinical vocabulary. Further details are provided in the Appendix.

We further linked each medication to its corresponding code in the World Health Organization’s Anatomical Therapeutic Chemical (ATC) classification system. This was achieved using the NHSBSA dm+d supplementary file^14^, which maps ATC codes to BNF chemical substance codes for products prescribed in Primary Care. The Dictionary of Medicines and Devices (dm+d) is a standardised resource containing descriptions and codes that represent medicines and medical devices used across the NHS. The dm+d and its associated files are distributed via the Terminology Reference Data Update Distribution (TRUD) service^15^, which is managed by NHS England and provides the delivery and licensing infrastructure for NHS terminologies, including the dm+d. These linkages were incorporated into the final knowledge base.

### Clinical validation

Clinical validation was carried out by selecting a random sample of unique medication-indication pairs (n = 645). To ensure a meaningful representation of the model’s performance across confidence levels. The sample was stratified by the LLM generated confidence score. Three clinical experts – two physicians and one clinical pharmacist – independently reviewed the sampled pairs, with each reviewer evaluating a third of the pairs. Each medication–indication pair was presented with its drug name and the indicated condition, and the reviewer was asked to judge whether that indication is a clinically appropriate use of the medication in practice. Reviewers applied their clinical knowledge and could consult standard reference sources such as the British National Formulary^7^, Summary of Product Characteristics, DrugBank^16^ and UpToDate^17^ for medication-indication pair verification. We provided guidelines that an indication should be marked correct if the medication is used to treat or prevent that condition in clinical practice (whether as a licensed indication or a well-recognised off-licence use), and incorrect if the medication is not used for that condition (e.g. an entirely unrelated or implausible indication). Each pair was thus assigned a binary outcome: 1 for clinically correct or 0 for incorrect. Ambgiouous pairs were resolved through discussion between the three clinicians. Using these ratings, we calculated the precision of the knowledge base as the proportion of true positives among the reviewed sample medication-indication pairs. Given the sample size of the internal validation sample, bootstrapped estimates of precision along with 95% confidence intervals were calculated.

### External validation

External validation with a real-world dataset (NHANES) was carried out to understand the similarity of the AI generated medicine-indication pairs to real world clinical practice. The final version of the medication-indication knowledge base had medications listed according to their BNF codes and descriptions, whilst the NHANES dataset uses the Multum Lexicon. Therefore, the first step was to harmonise the drug names between the two data sources. This was performed using approximate string matching, as described in the knowledge base creation section. Common American names for medications were changed to their British names first (e.g. acetaminophen to paracetamol) to facilitate the matching process.

Validation was carried out on a random subset of 250 medication-indication pairs from the NHANES data by the same individuals who conducted internal validation, with each checker assigned roughly one-third of the medication-indication pairs. The random subset was manually compared against the medication-indication pairs generated by GPT4 to score the similarity of the GPT4 generated pairs to the reported indication of the medicine in NHANES. A binary scoring system was used, such that a score of 1 was assigned where the medication-indication pair was present in both the knowledge base and the NHANES dataset, and a score of 0 was assigned where the indication reported in NHANES was not present in the knowledge base. As with the internal validation, ambiguous cases were resolved by the three clinicians through discussion. Overall agreement between our knowledge base and the pre-pandemic NHANES dataset was calculated as the proportion of the medication-indication pairs from NHANES that were correctly classified in the GPT4 generated knowledge base.

## Results

A total of 1,540 unique chemical entities were extracted from the EPD and included in the final prompt pipeline. This resulted in 10,853 medication-indication pairs. Examples of the outputs generated are shown in Table 1. We observed that many of the outputs were clinically correct but needed additional data cleaning and standardisation for several reasons: inconsistent use of acronyms and abbreviations (e.g. “HIV infection” versus “Human immunodeficiency virus infection”), spelling mistakes, inconsistent use of capitalisation and plurals, and duplications due to clinically synonymous terms (e.g. “eczema” versus “atopic dermatitis”).

**Table 1:** Examples of medication-indication pairs that were clinically correct but unstandardised, or clinically incorrect.

| Type of indication | Example |  |
| --- | --- | --- |
|  | Medication | Indication(s) |
| Clinically correct, but inconsistent terminology | Alendronic acid/colecalciferol | Paget’s disease, Paget’s disease of the bone |
|  | Benozoyl peroxide | Acne, Acne vulgaris |
|  | Exemestame | Advanced breast cancer, Advanced-stage breast cancer |
| Clinically incorrect indication | Co-tenidone | Cardiac arrhythmias |
|  | Metformin hydrochloride/pioglitazone | Prediabetes |
|  | Alprazolam | Depression |

### Clinical validation

In the random sample of medicine-indication pairs included in the clinical validation set (n = 645), most responses were assessed to be clinically correct by the checking physician or pharmacist, with an overall bootstrapped precision of 83.2% (95% CI 80.3% to 86.0%). We analysed the relationship between GPT-4’s self-generated confidence scores and the clinical validity of its medication–indication outputs (Figure 1). Lower confidence scores strongly associated with a higher error rate. For example, medication–indication pairs with confidence scores below 0.7 showed a markedly higher proportion of clinically implausible or incorrect associations, while those ≥ 0.75 had substantially fewer errors.

**Figure 1:**
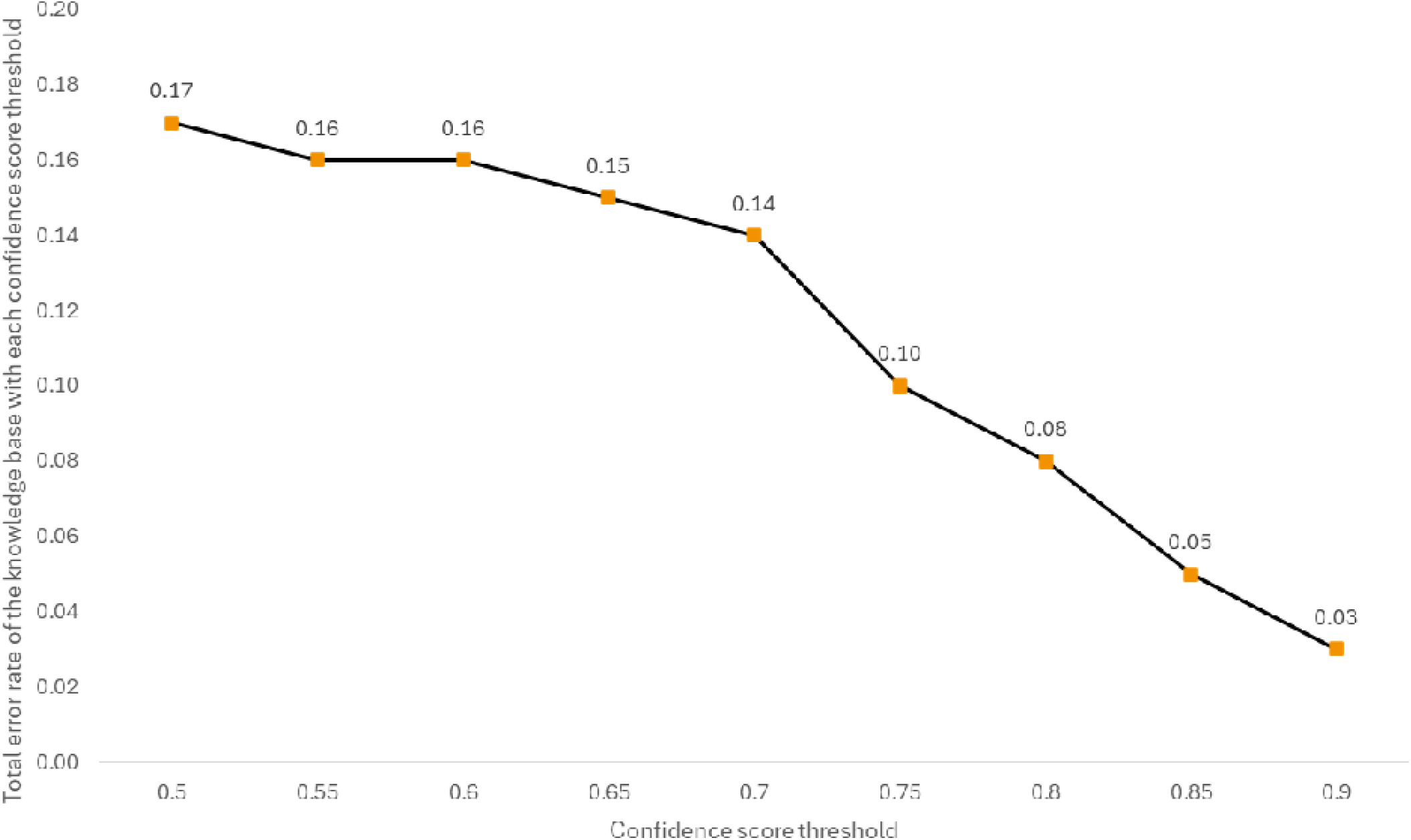
Graph of total error rates of the knowledge base using each confidence score threshold.

We further examined the correct and incorrect medication-indication pairs to see if there were any patterns in the indications that were in each category. Table 2 contains the top 10 indications by frequency among correct and incorrect medication-indication pairs. The indications appearing at high frequencies are similar in both correct and incorrect pairs – e.g., migraine is the most frequent indication in both, while pneumonia and depression are in the top 5 in both

**Table 2:** Frequencies of indications in the correct and incorrect medication-indication pairs from clinical validation.

| Type of medication/indication pair | Indication | Frequency (%) |
| --- | --- | --- |
| Incorrect | Migraine | 16 (15) |
|  | Breast cancer | 14 (13) |
|  | Depression | 13 (12) |
|  | Systemic lupus erythematosus | 8 (7) |
|  | Pneumonia | 8 (7) |
|  | Angina | 7 (6) |
|  | Liver cirrhosis | 7 (6) |
|  | Asthma | 6 (6) |
|  | Psoriatic arthritis | 4 (4) |
|  | Stroke | 4 (4) |
| Correct | Migraine | 60 (11.2) |
|  | Angina | 56 (10.4) |
|  | Pneumonia | 44 (8.2) |
|  | Stroke | 36 (6.7) |
|  | Depression | 35 (6.5) |
|  | Asthma | 31 (5.8) |
|  | Breast cancer | 25 (4.7) |
|  | Systemic lupus erythematosus | 23 (4.3) |
|  | Discoid lupus erythematosus | 17 (3.2) |
|  | Psoriatic arthritis | 11 (2.0) |

We then investigated the types of errors at different confidence score thresholds. Table 3 contains examples of these. Incorrect indications were returned at all confidence levels, but the frequency of pairs where the returned indication was closely related to the actual indication, or where the actual treatment was used was related to the medicine in the medication-indication pair was higher at the higher confidence scores.

**Table 3:** Examples of errors arising in the clinical validation.

| Type of error | Medication-indication pair example | Reason for evaluation | Confidence score |
| --- | --- | --- | --- |
| Incorrect indication | Omega-3 marine triglycerides – Depression | Not a typical treatment for depression | 0.5 |
|  | Terpin hydrate – Pneumonia | A mucolytic; not enough evidence supporting their use in pneumonia | 0.5 |
|  | Naftidrofuryl oxalate – Stroke | Used to treat peripheral vascular disorders | 0.6 |
|  | Simple (linctus) – Angina | Simple linctus is for treating<br>coughs | 0.85 |
| Incorrect but closely<br>related to actual<br>medication/indication | Testosterone – Breast cancer | Only specific<br>testosterones<br>are used to<br>treat breast<br>cancer | 0.75 |
|  | Formoterol/glycopyrronium/budesonide – Asthma | Used for COPD | 0.85 |
|  | Prasterone – Lupus | Steroids are<br>used for lupus,<br>but not<br>prasterone<br>specifically | 0.9 |

As observed in Figure 1, there was a relatively large decrease in error rate between confidence score thresholds of 0.7 and 0.75. In the validation sample, 465 medication indication pairs had confidence scores of at least 0.75, of which 418 were assessed to be correct. This corresponds to a bootstrapped precision of 89.85%, 95% CI 87.10% to 92.47%. To balance the number of rows in the knowledge base and precision, we decided to limit our knowledge base to medication indication pairs with confidence scores of 0.75 and higher, resulting in 8,653 medication indication pairs. We used this filtered knowledge base for external validation.

Finally, we examined the error rates by ICD-10 chapter to investigate whether there were any systematic differences between indication type. Table 4 displays the percentage of correct medication-indication pairs by ICD-10 chapter, where there were more than a total of 5 pairs being checked within that chapter. The proportion of clinically accurate pairs ranged from 65% for neoplasms, to 100% for diseases of the genitourinary system, and endocrine, nutritional and metabolic diseases (though note that these were based on only 7 and 10 pairs respectively).

**Table 4:** Number and proportion of clinically accurate medication-indication pairs by ICD-10 chapter.

| ICD 10 chapter | Number of correct pairs | Number of pairs checked | Percentage of correct pairs |
| --- | --- | --- | --- |
| Endocrine, nutritional and metabolic diseases | 10 | 10 | 100 |
| Diseases of the genitourinary system | 7 | 7 | 100 |
| Certain infectious and parasitic diseases | 62 | 65 | 95.38 |
| Diseases of the circulatory system | 111 | 126 | 88.1 |
| Diseases of the musculoskeletal system and connective tissue | 30 | 35 | 85.71 |
| Symptoms, signs and abnormal clinical and laboratory findings, not elsewhere classified | 6 | 7 | 85.71 |
| Diseases of the nervous system | 86 | 102 | 84.31 |
| Diseases of the respiratory system | 76 | 92 | 82.61 |
| Diseases of the skin and subcutaneous tissue | 37 | 45 | 82.22 |
| Diseases of the digestive system | 26 | 34 | 76.47 |
| Mental and behavioural disorders | 39 | 54 | 72.22 |
| Neoplasms | 26 | 40 | 65 |

### External validation

A total of 2,899 unique medication-indication pairs in NHANES had matched medication names in the knowledge base. A random sample of 250 (8.6%) were extracted for validation. Of these, 210 were captured in the LLM generated knowledge base, resulting in an overall agreement of 84% between the two sources. Among the 40 medication-indication pairs not captured by the knowledge base, only 2 (5%) were valid indications that were missed by the knowledge base, 7 (17.5%) were missed off-licence indications and the majority-31 (77.5%) indications-were considered to be likely to be incorrectly reported by patients (See Table 5 for examples).

**Table 5:**
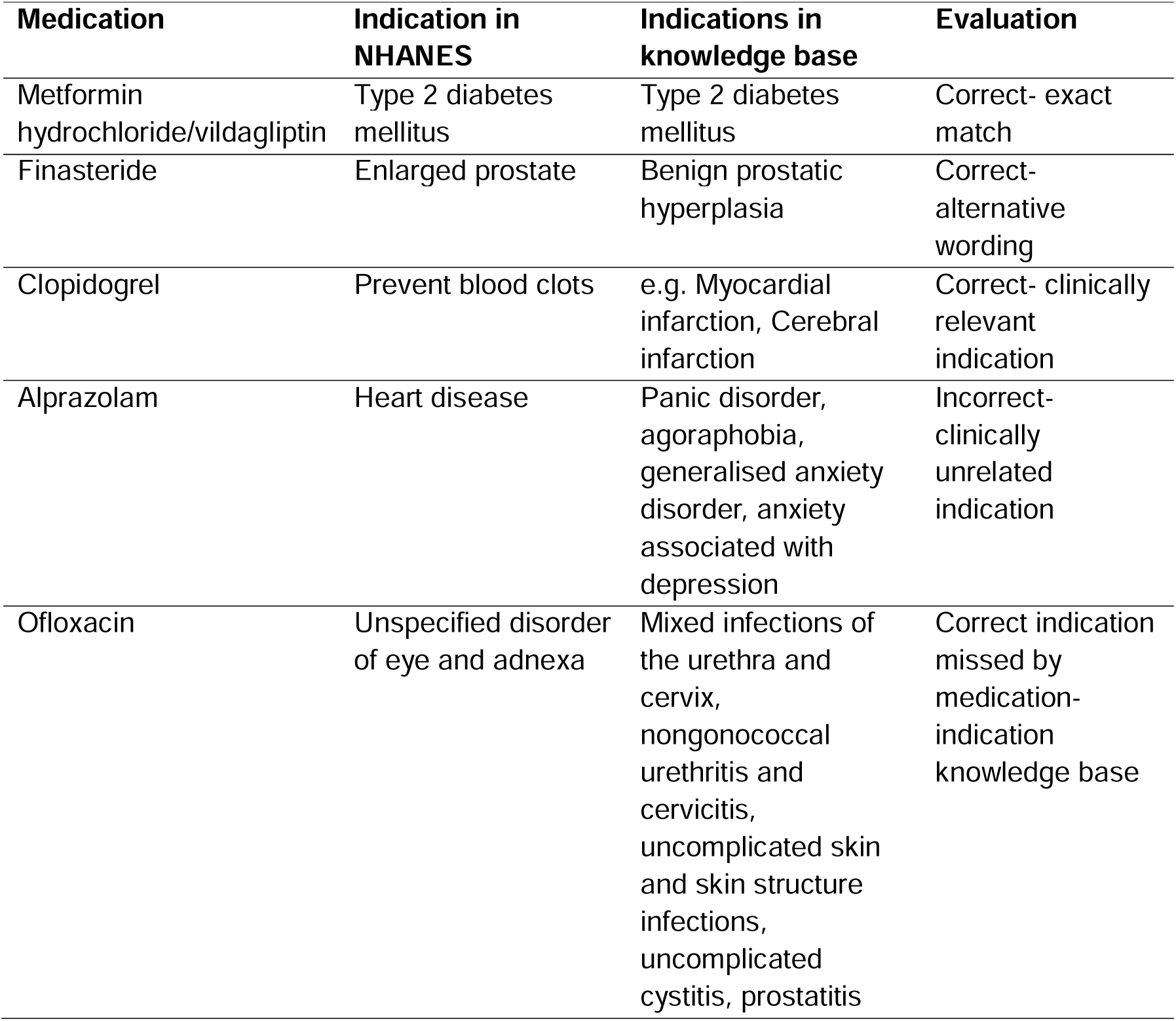
Examples from the external validation error analysis.

The most recent version of the medication-indication database contains BNF chapters, sections and paragraphs and ingredient-level BNF codes along with ATC codes for each medication and ICD10 and SNOMED CT codes for indications. Examples of the full set of information available in the knowledge base is presented in Table 6.

**Table 6:** Examples of medication-indication pairs in the knowledge base including information about BNF chapters, sections and paragraphs as well as the purpose of treatment

| BNF Chapter |  | BNF Section |  | BNF Paragraph |  | BNF Chemical Substance |  | ATC Code | Indication | Purpose |
| --- | --- | --- | --- | --- | --- | --- | --- | --- | --- | --- |
| Code | Name | Code | Name | Code | Name | Code | Name |  |  |  |
| 3 | Respiratory System | 305 | Antihistamines, hyposensitisation and allergic emergencies | 30401 | Antihistamines | 0304010A0 | Acrivastine | R01BA52 | Hay fever | Treatment |
| 11 | Eye | 1106 | Treatment of glaucoma | 110600 | Treatment of glaucoma | 1106000AF | Bimatoprost | S01EE03 | Primary open-angle glaucoma | Treatment |
| 2 | Cardiovascular system | 208 | Anticoagulants and protamine | 20801 | Parenteral anticoagulants | 0208010K0 | Heparin sodium | B01AB01 | Pulmonary embolism | Treatment |
| 6 | Endocrine system | 601 | Drugs used in diabetes | 60102 | Antidiabetic drugs | 0601022B0 | Metformin hydrochloride | A10BA02 | Type 2 diabetes | Treatment |
| 13 | Skin | 1311 | Skin cleaners, antiseptics and desloughing agents | 1131105 | Phenolics | 1311050K0 | Hexachlorophene | D08AE01 | Surgical site infection | Prevention |
| 4 | Central Nervous System | 402 | Drugs used in psychoses and related disorders | 40201 | Antipsychotic drugs | 0402010P0 | Pericyazine | N05AC01 | Acute psychosis | Treatment |
| 8 | Malignant disease and Immunosuppression | 802 | Drugs affecting the immune response | 80202 | Corticosteroids and other immunosuppressants | 0802020U0 | Sirolimus | L04AH01 | Organ transplant rejection | Prevention |

## Discussion

Using a combination of general-purpose foundation LLM (GPT-4) and a health data specific model optimised for natural language processing of electronic health records, we have developed a medication-indication knowledge base suitable for use in health data analytics applications and pharmacoepidemiological research. The knowledge base had an accuracy similar to previously developed medicines knowledge databases, comparable to those created using extensive expert human involvement^2^, whilst covering all medicines dispensed by pharmacies in the United Kingdom.

Our analysis confirmed a strong relationship between GPT-4’s self-generated confidence scores and clinical validity. While not probabilistic, these scores proved useful as a heuristic for filtering unreliable outputs - lower scores were associated with more frequent errors, while pairs scoring ≥ 0.75 demonstrated notably higher precision. This observation is consistent with prior studies showing that LLM-generated confidence, often derived from token-level likelihoods or embedding similarity, can correlate with factual accuracy - particularly when elicited through structured prompts^18^. Though uncalibrated, such scores have been shown to reflect answer reliability in both general and medical contexts^19^. Given LLMs’ known tendency for overconfidence, confidence-based filtering represents a practical safeguard^20^. However, these scores should not be interpreted as formal probabilities.

Previous approaches to developing medication-indication knowledge bases have used a spectrum of approaches from fully data driven to fully manual creation. For example, the MEDication Indication (MEDI) database^21^ contains medication-indication pairs from four public medication resources: RxNorm, SIDER 2, MedlinePlus, and Wikipedia. Natural language processing (NLP) and ontology relationships were used to extract indications for 3,112 single-ingredient medications. resulting in 63,343 medication-indication pairs. It was subsequently used for applications such as drug-repurposing, validation of other knowledge bases, relation extraction from clinical text and the identification of adverse drug events (Wei et al. 2021). A second version of MEDI (MEDI-2) was developed recently^22^ to reflect more timely medication-indication information, in which information was extracted from the original four sources, as well as two additional sources: MayoClinic and SIDER 4.1. Although a more extensive range of sources is required for broader coverage, the extraction and mapping procedure may not be feasible with a very large number of sources. MEDI and MEDI-2 do not differentiate between whether a medication treats a particular indication or prevents it; depending on the use-case, users may be interested only in treatment or prevention-in which case errors may be introduced by the lack of distinction between both. The AI-assisted knowledge base addresses several of these limitations, by capturing off-licence indications and distinguishing between preventative and treatment relationships.

A combination of automatic and manual methods was used to develop the LabeledIn^2^ database. Automated extraction and tagging of disease and indication mentions from drug labels was followed by manual evaluation by clinical experts to produce a final gold-standard. LabeledIn is much smaller than MEDI and MEDI-2 and only catalogues 250 most frequently highly accessed drugs in PubMed Health. The authors attempted to scale this by outsourcing the manual evaluation to unknown workers through the Amazon Mechanical Turk technical environment and found that this resulted in high accuracy on a set of control medication-indication pairs (those known to be correct) in a relatively short span of time^23^. Moodley et al.^24^ followed a similar approach in the development of their medication-indication knowledge base called InContext-they used automatic methods to detect mentions of diseases, but relied on human annotation to select mentions of indications from other non-indication disease mentions. They additionally included contextual information that also appears in SPLs, including co-prescribed medication, co-therapies, co-morbidities, genetics and temporal information.

Both InContext and LabeledIn rely on drug labels only, increasing the chances of the omission of off-licence indications. The named entity recognition tools themselves were shown to omit some indications from the labels that had to be manually added by clinical experts to MEDI-2. Additionally, medication-indication information may change over time as new medicines are developed, or existing medications are approved for newer indications. Repeating manual annotation processes in the longer term to reflect these changes may not be feasible, given the associated costs and resources utilised.

This knowledge base could be used in computational drug repurposing, as a reference dataset of known medication-indication relationships required to identify and predict novel associations. A gold-standard dataset created by Gottlieb et al.^25^ has widely been used for this purpose^26,27^. It contains 1933 medication-indication pairs for 593 drugs from DrugBank and 313 diseases from the Online Mendelian Inheritance in Man (OMIM) database. Our AI-assisted medication-indication knowledge contains many more medication-indication pairs, so is likely to have broader coverage of licensed and off-licence indications-which is necessary to filter truly novel predictions. The knowledge base could similarly be used in the extraction or identification of adverse drug events, to serve the purpose of filtering out indications-for example, Kuhn et al. developed SIDER^3^, an adverse drug event database which also includes known indications which were used to remove false positive adverse events. However, the known indications were extracted from drug package inserts-using a source with information on off-licence indications would provide a more robust filter to remove clinically valid but off-label indications.

Our knowledge base is generated by a large language model and so may contain some inaccuracies. We developed an extensive error detection and data cleaning pipeline to tackle these and would advise against using the raw output from GPT-4 for indication mapping. Although the accuracy of the knowledge base was comparable to existing manually created data sources, the proportion of detected errors means that the knowledge base should not be used in clinical practice.

The knowledge base is not exhaustive and does not contain all medications and all their licensed and off-licence indications-although the aim was for the knowledge base to be as comprehensive as possible whilst maintaining highest levels of accuracy. The clinical validation process focused on evaluating the indications that were present in the knowledge base, and not on detecting which medications or indications were omitted. Similarly, the external validation process only included medications that were reported in both NHANES and the knowledge base. Approximate string matching of the medication names to BNF descriptions may not have been fully accurate, as the aim of the algorithm used was to find the most similar string possible. This may not be suitable for all medication names, given the sometimes-subtle differences in spelling between completely different medications. Finally, the timeliness of the knowledge base is limited by the period of the trained knowledge of the GPT-4 LLM.

Further development of this AI-assisted knowledge base is ongoing, including the inclusion of medicines used in inpatient hospital settings; additional cleaning and harmonisation of indication names is being conducted using the MedCAT annotation tool^13^. Future studies should benchmark the performance of GPT-4 against open-source LLMs such as DeepSeek^28^ or LLAMA^29^ – at the time of this study, GPT-4 was the best in class, with no equivalent alternatives. Benchmarking may also be carried out with different versions of GPT-4 that have since been introduced, such as GPT-4 Turbo, GPT-4o and o1 to name a few^30^.

## Conclusion

Given appropriate prompt engineering and output processing, LLMs can be used to generate information about the clinical indications of medicines with sufficient accuracy to be used in research and analytics applications. The approach overcomes several limitations of existing approaches, and is much more scalable and maintainable than manual creation by human experts. The resulting knowledge base is machine readable, uses standard terminologies and is annotated with widely used medicine and clinical coding schemes, making it well-suited to support real-world evidence and pharmacoepidemiological research, computational drug repurposing and adverse drug event detection.

## Data Availability

All data sources used in this study are publicly available:
GPT4- https://openai.com/index/gpt-4/
MedCAT- https://github.com/CogStack/MedCAT
Dictionary of medicines and devices (dm+d)-Dictionary of medicines and devices (dm+d) | NHSBSA
EPD- https://opendata.nhsbsa.net/dataset/english-prescribing-data-epd

https://openai.com/index/gpt-4/

https://github.com/CogStack/MedCAT

https://www.nhsbsa.nhs.uk/pharmacies-gp-practices-and-appliance-contractors/dictionary-medicines-and-devices-dmd

https://opendata.nhsbsa.net/dataset/english-prescribing-data-epd

## Ethics approval and consent to participate

All data sources used in this study are publicly available:

- GPT4- https://openai.com/index/gpt-4/
- MedCAT- https://github.com/CogStack/MedCAT
- Dictionary of medicines and devices (dm+d)-Dictionary of medicines and devices <u>(dm+d) | NHSBSA</u>
- EPD- https://opendata.nhsbsa.net/dataset/english-prescribing-data-epd

## Consent for publication

Not applicable

## Availability of data and materials

Not applicable

## Competing interests

The authors declare that they have no competing interests

## Funding

Not applicable

## Authors’ contributions

EB, MF, AM, LZ and BB finetuned the prompt pipeline for GPT-4 and created the output containing the medication-indication pairs for the knowledge base. They also carried out the harmonisation of indication names. AB, FM and BB conducted the clinical validation and SD, FM and AB conducted the external validation for the AI-assisted knowledge base. BB and SD were the major contributors in writing the manuscript. All authors read and approved the final manuscript.

## Acknowledgements

We thank Alex Castanon, Godspower Oboli and Luisamanda Selle Arocha for their work on the clinical validation of the knowledge base.

## Appendix

Prompt used for GPT-4 to generate indications using medications from the English Prescribing

Dataset.

Model: GPT4, via api

Plugins: None

You are a medically trained epidemiologist and will be generating a table to describe the clinical indications for medicines. The table should have the following columns: “Medicine name”, “Indication”, “Purpose”, “Medicine Class”, “Confidence score”. For purpose, indicate if the medicine is for prevention or treatment. For confidence score, return a score in the range 0 to 1 which reflects the level of certainty that the indication is clinically valid, were 0 represents the lowest certainty and 1 represents highest certainty. Provide the sources used to generate the confidence scores, and the principles used in quantifying confidence as well as a detailed breakdown of all thresholds for confidence scores. Indication should be disease name rather than a group of diseases. The table should have one row for each indication. Use medical terminology throughout and British English spellings. Make the list comprehensive and do not limit the number of indications given but it must include only approved indications. Do this for prednisolone as an example.”

Preliminary list of chemical substances (drugs) to test:

Atorvastatin

Levothyroxine Sodium

Omeprazole

Amlodipine

Ramipril

Bisoprolol Fumarate

Cholecalciferol

Metformin Hydrochloride

Prednisolone

### Methodology provided by GPT-4 for the calculation of confidence scores

“Sources used to generate the confidence scores include clinical trials, systematic reviews, and meta-analyses from reputable databases such as PubMed, Cochrane, and ClinicalTrials.gov.

The principles used in quantifying confidence include the quality of the evidence (randomised controlled trials provide the highest level of evidence), the consistency of the results across different studies, and the directness of the evidence (whether the studies directly compare the medicine to a placebo or to another treatment).

The thresholds for confidence scores are as follows:

- 0.8 to 1.0: High confidence, based on multiple high-quality studies with consistent results.
- 0.6 to 0.79: Moderate confidence, based on one high-quality study or multiple studies with some limitations.
- 0.4 to 0.59: Low confidence, based on studies with significant limitations.
- 0.0 to 0.39: Very low confidence, based on expert opinion or case reports.

Please note that this is a simplified example and the actual process of generating confidence scores is more complex and involves a detailed assessment of the evidence.”

### Use of MedCAT for indication cleaning

MedCAT (Medical Concept Annotation Toolkit) is a natural language model trained on hospital electronic medical records, specifically designed for natural language processing of clinical text. It uses a self-supervised machine learning algorithm to extract concepts using vocabularies such as UMLS and SNOMED-CT. There are four public models available to use, of which we used the SNOMED international model (full SNOMED modelpack trained on MIMIC-III) downloaded using the medcat library in Python.

We created a preliminary indication cleaning pipeline in Python, which included steps such as reconciling discrepancies in the use of hyphens for the same term (e.g. “non-valvular” vs. “nonvalvular”, “early-stage breast cancer” vs. “early stage breast cancer”), singular and plural forms of the same term (e.g. “infection” vs. “infections”), abbreviations and spelling inconsistencies between UK and US spellings (e.g. “tumour” vs. “tumor”). We then used the get.entities() function from the medcat library to extract SNOMED codes and ICD-10 codes in separate columns for the indication in each row of the knowledge base. Since several codes for the same indication were possible, we extracted the first code extracted for each indication, as MedCAT orders these by level of similarity with the relevant SNOMED concept. The generated codes underwent validation by clinical experts, who ascertained the validity of a subset of indications at the ICD-10 three-character code level.

This was an iterative process – preprocessing steps were added based on the quality of the output of the previous iteration to maximise the number of code matches. For example, after running the steps highlighted above it became apparent that some indications were too specific to be matched to a specific SNOMED or ICD-10 code. These were then grouped into slightly broader categories before rerunning the entire pipeline – for example, “seasonal allergic rhinitis”, “perennial allergic rhinitis”, “hay fever” and “pollen-induced allergic rhinitis” were all changed to the term “allergic rhinitis”. Once this process was complete, the generated codes underwent validation by clinical experts, who evaluated the suitability of the ICD-10 codes generated for a subset of the indications using the descriptions provided in the WHO ICD-10 browser (available here: https://icd.who.int/browse10/2019/en). Validation was carried out by two clinicians, with ambiguous terms being resolved by a third clinician/

## Notes

### Competing Interest Statement

The authors have declared no competing interest.

### Funding Statement

This study did not receive any funding

## References

1. Zhu Y, Elemento O, Pathak J, Wang F. Drug knowledge bases and their applications in biomedical informatics research. Brief Bioinform. 2019;20(4):1308–1321. doi:10.1093/bib/bbx169

2. Khare R, Li J, Lu Z. LabeledIn: Cataloging Labeled Indications for Human Drugs. J Biomed Inform. 2014;52:448–456. doi:10.1016/j.jbi.2014.08.004

3. Kuhn M, Letunic I, Jensen LJ, Bork P. The SIDER database of drugs and side effects. Nucleic Acids Res. 2016;44(D1):D1075–D1079. doi:10.1093/nar/gkv1075

4. Bhatt A, Roberts R, Chen X, et al. DICE: A Drug Indication Classification and Encyclopedia for AI-Based Indication Extraction. Front Artif Intell. 2021;4. doi:10.3389/frai.2021.711467

5. InContext: curation of medical context for drug indications | Journal of Biomedical Semantics | Full Text. Accessed April 15, 2025. https://jbiomedsem.biomedcentral.com/articles/10.1186/s13326-021-00234-4

6. Jung K, LePendu P, Chen WS, et al. Automated Detection of Off-Label Drug Use. Sarkar IN, ed. PLoS ONE. 2014;9(2):e89324. doi:10.1371/journal.pone.0089324

7. BNF content published by NICE. April 10, 2025. Accessed April 15, 2025. https://bnf.nice.org.uk/

8. Van Veen D, Van Uden C, Blankemeier L, et al. Adapted large language models can outperform medical experts in clinical text summarization. Nat Med. 2024;30(4):1134–1142. doi:10.1038/s41591-024-02855-5

9. English prescribing data (EPD) | NHSBSA. Accessed April 15, 2025. https://www.nhsbsa.nhs.uk/prescription-data/prescribing-data/english-prescribing-data-epd

10. NHANES Questionnaires, Datasets, and Related Documentation. Accessed April 15, 2025. https://wwwn.cdc.gov/nchs/nhanes/continuousnhanes/default.aspx?Cycle=2017-2020

11. Achiam J, Adler S, Agarwal S, et al. GPT-4 Technical Report. 10.48550/arXiv.2303.08774

12. Sariyar M, Borg A. The RecordLinkage Package: Detecting Errors in Data. R J. 2010;2(2):61–67. doi:10.32614/RJ-2010-017

13. Kraljevic Z, Searle T, Shek A, et al. Multi-domain clinical natural language processing with MedCAT: The Medical Concept Annotation Toolkit. Artif Intell Med. 2021;117:102083. 10.1016/j.artmed.2021.102083

14. Dictionary of medicines and devices (dm+d) | NHSBSA. Accessed April 15, 2025. https://www.nhsbsa.nhs.uk/pharmacies-gp-practices-and-appliance-contractors/dictionary-medicines-and-devices-dmd

15. Log in - TRUD. Accessed April 15, 2025. https://isd.digital.nhs.uk/trud/users/guest/filters/0/login/form

16. Knox C, Wilson M, Klinger CM, et al. DrugBank 6.0: the DrugBank Knowledgebase for 2024. Nucleic Acids Res. 2024;52(D1):D1265–D1275. doi:10.1093/nar/gkad976

17. Sign in - UpToDate. Accessed April 15, 2025. https://www.uptodate.com/login

18. Kadavath S, Conerly T, Askell A, et al. Language Models (Mostly) Know What They Know. Published online November 21, 2022. doi:10.48550/arXiv.2207.05221

19. Tian K, Mitchell E, Zhou A, et al. Just Ask for Calibration: Strategies for Eliciting Calibrated Confidence Scores from Language Models Fine-Tuned with Human Feedback. In: Bouamor H, Pino J, Bali K, eds. Proceedings of the 2023 Conference on Empirical Methods in Natural Language Processing. Association for Computational Linguistics; 2023:5433–5442. doi:10.18653/v1/2023.emnlp-main.330

20. Chhikara P. Mind the Confidence Gap: Overconfidence, Calibration, and Distractor Effects in Large Language Models. Published online February 16, 2025. doi:10.48550/arXiv.2502.11028

21. Wei WQ, Cronin RM, Xu H, Lasko TA, Bastarache L, Denny JC. Development and evaluation of an ensemble resource linking medications to their indications. J Am Med Inform Assoc JAMIA. 2013;20(5):954–961. doi:10.1136/amiajnl-2012-001431

22. Zheng NS, Kerchberger VE, Borza VA, Eken HN, Smith JC, Wei WQ. An updated, computable MEDication-Indication resource for biomedical research. Sci Rep. 2021;11(1):18953. doi:10.1038/s41598-021-98579-4

23. Khare R, Burger JD, Aberdeen JS, et al. Scaling drug indication curation through crowdsourcing. Database. 2015;2015:bav016. doi:10.1093/database/bav016

24. Moodley K, Rieswijk L, Oprea T, Dumontier M. InContext: curation of medical context for drug indications. J Biomed Semant. 2021;12. 10.1186/s13326-021-00234-4

25. Gottlieb A, Stein GY, Ruppin E, Sharan R. PREDICT: a method for inferring novel drug indications with application to personalized medicine. Mol Syst Biol. 2011;7(1):496. doi:10.1038/msb.2011.26

26. Jiang HJ, Huang YA, You ZH. Predicting Drug-Disease Associations via Using Gaussian Interaction Profile and Kernel-Based Autoencoder. BioMed Res Int. 2019;2019(1):2426958. doi:10.1155/2019/2426958

27. Zhang W, Yue X, Lin W, et al. Predicting drug-disease associations by using similarity constrained matrix factorization. BMC Bioinformatics. 2018;19(1):233. doi:10.1186/s12859-018-2220-4

28. DeepSeek-AI, Liu A, Feng B, et al. DeepSeek-V3 Technical Report. Published online February 18, 2025. doi:10.48550/arXiv.2412.19437

29. Grattafiori A, Dubey A, Jauhri A, et al. The Llama 3 Herd of Models. Published online November 23, 2024. doi:10.48550/arXiv.2407.21783

30. Models - OpenAI API. Accessed April 15, 2025. https://platform.openai.com

